# THE ERA OF CORONAVIRUS; KNOWLEDGE, ATTITUDE, PRACTICES, AND BARRIERS TO HAND HYGIENE AMONG MAKERERE UNIVERSITY STUDENTS AND KATANGA COMMUNITY RESIDENTS

**DOI:** 10.1101/2020.06.05.20123042

**Authors:** Julius Nuwagaba, Dave Darshit Ashok, Thomas Balizzakiwa, Ibrahim Kisengula, Edna Joyce Nagaddya, Meddy Rutayisire

## Abstract

**Background:** China reported the Novel Coronavirus at the end of the year 2019 which was, later on, declared a Pandemic by the WHO. Proper hand hygiene was identified as one of the simplest most cost-effective Covid-19 control and prevention measures. It is therefore very important to understand the compliance of the community to hand hygiene.

**Method:** A descriptive cross-sectional study was conducted among the undergraduate students of Makerere University and residents of Katanga slum from 17th to 22nd of March, 2020. An interviewer guided questionnaire with questions on knowledge, attitude, practice, and barriers to hand hygiene was used in data collection. The collected data was analyzed using Microsoft office excel 2016 and STATA 15 software. A 95% confidence interval was used and statistical significance was P<0.05.

**Results:** Only 8.4% of the participants had good knowledge of hand hygiene. 11.7% of the university students had good knowledge compared to 0.9% of the Katanga residents. 29.0% of the participants had a good attitude while 50.1% had a moderate attitude to hand hygiene. University students were 6.3 times (OR: 6.3, 95%C1: (2.1 – 18.5), P=0.001) more likely to have good knowledge while Katanga residents were 3.6 times (OR: 3.6, 95%C1: (1.5 – 8.4), P=0.003) more likely to have good attitude to hand hygiene. Only 19.6% accomplished all the seven steps of handwashing. 38.4% of the participants still greeted by handshaking and 60.1% noted lack of soap as a barrier to hand hygiene and 62.9% reported having more than three barriers to hand hygiene. Participants that had been taught handwashing were more likely to have better hand hygiene knowledge and practice.

**Conclusion:** Despite a fair attitude, deficiency of knowledge coupled with many barriers such as Lack of soap hindered the Practice of proper hand hygiene. Public health involvement to promote hand hygiene must be promoted.

## BACKGROUND

Coronaviruses (CoV) are a large family of zoonotic viruses that cause illness that ranges from the common cold to more severe diseases such as Middle East Respiratory Syndrome (MERS-CoV) and Severe Acute Respiratory Syndrome (SARS-CoV) (1,2). A novel coronavirus (nCoV) is a new strain that had not been previously identified in humans until the end of 2019 in Wuhan, Hubei province, China (3,4). The 2019-nCoV can transmit among humans (5,6) and as of 29^th^ May 2020, there were 5,701,337 cases and 357,688 deaths globally (7). Among other forms of spread, a person can get COVID-19 by touching a surface or object that has the virus on it and then touching their mouth, nose, or possibly their eyes (8,9). Handwashing with soap can reduce the risk of acute respiratory infections by 16% to 23% (10). WHO and the Uganda Ministry of Health recommend Hand hygiene as one of the essential means to prevent the spread of all infections and in particular COVID 19. Other measures recommended include maintaining social distance, avoiding crowds, practicing respiratory hygiene, avoiding touching eyes, nose, and mouth, keeping up to date on the latest information from trusted sources, self-quarantine, cleaning frequently touched surfaces, and seeking medical care in case of symptoms (11,12). The promotion of safe hygiene is the single most cost-effective means of preventing infectious disease (13). During a global pandemic, one of the cheapest, easiest, and most important ways to prevent the spread of a virus is to wash your hands frequently with soap and water (14–16).

The promotion of hand hygiene behavior is a complex issue (17,18). Reasons for non-compliance with recommendations occur at individual, group, and institutional levels (19). Individual factors such as social cognitive and psychological determinants (i.e. knowledge, attitude, intentions, beliefs, and perceptions) provide additional insight into hand hygiene behavior (20). Perceived barriers to adherence to hand hygiene practice recommendations include inaccessible hand hygiene supplies, forgetfulness, lack of knowledge of guidelines, insufficient time for hand hygiene (21). Despite considerable efforts, compliance with hand hygiene as a simple infection-control measure remains low (22) and hygiene is suboptimal in both community and healthcare settings in African countries (23).

Several studies have compared different hand hygiene methods in hospital settings (21). In contrast, few studies have been published on the effect of hand hygiene on bacterial contamination of hands in the community(24,25). Makerere University, the largest university in Uganda is one of the high-risk areas of COVID-19 transmission due to factors like a large student and staff community (26). The university is surrounded by several communities including Katanga slum which is located between the main campus and the Medical school. These are high concentration areas with a high risk of community transmission of COVID 19. This research served to identify gaps in the knowledge, attitude, and practices and barriers regarding hand-hygiene among the Makerere University students and Katanga slum residents. The results from this study are very useful in paving a way for comprehensive intervention for successful behavior change programs on measures for the implementation of proper hand hygiene.

## METHOD

### Study design and setting

We employed a descriptive cross-sectional study design among the Makerere University medical students and non-medical students residing in halls of residence. Data was also collected from the residents of Katanga slum, a settlement located in the valley between Mulago Hospital and Makerere University and its map can be accessed on https://goo.gl/maps/fqMmkk6cR1k4pNVbA. The study included only undergraduate students and Katanga slum residents aged 18 years and above who were able to understand English or Luganda languages.

### Data collection

Data were collected from 17th to 22 March 2020 using an interviewer-administered structured questionnaire. The original English questionnaire was also translated into Luganda, the local language spoken by residents of the Katanga community. Before using the tool, the Luganda tool was translated back to English to check for consistency. Data were collected on sociodemographic characteristics, knowledge, attitude, and practice of hand hygiene, and barriers to proper hand hygiene.

### Data analysis

Data was entered using epicollect5 software. This was after a thorough check for completion. The data were exported and analyzed using Microsoft office excel 2016 and STATA 15 software. Frequency distribution and percentages were used to analyze data in univariate, bivariate, and multivariate analysis. 9 parameters were used to assess the knowledge of patients on hand hygiene. Participants who got 8 to 9, 6 to 7, and below 6 correct answers were taken to have good, moderate, and poor knowledge respectively. To measure attitude to hand hygiene, a 5 point Likert scale analysis was used to strongly agree with 5, Agree with a 4, Neutral with a 3, Disagree with a 2, and strongly disagree with a 1. 4 parameters were used to assess the attitude to hand hygiene (total score of 20). Bloom’s cut-off of 80% was used to determine the attitude of the participants >= 80% (16) and above, 60%>= X<80% (12 to 15) and <60% (11 and below) were taken to have good, moderate and poor attitudes respectively. The practice of participants on hand hygiene was assessed based on how they greet in the era of COVID 19 and their ability to demonstrate the 7 steps of handwashing. The barriers of participants to hand hygiene were assessed on 6 parameters

Association between participants’ knowledge, attitude, practice, and barriers was represented in odds ratio with a 95% confidence interval using multivariate analysis. For all tests conducted in this study, a statistically significant Level was accepted at p<0.05. Spearman’s coefficient correlation was used to assess the relationship between knowledge and attitude to hand hygiene. The data set can be accessed via a link provided in the Supplementary Materials section.

### Ethical Considerations

Ethical approval to conduct the study was obtained from the Mulago Hospital Research and Ethics Committee. The approval to conduct the study within Katanga Slum was obtained from the Chairpersons of both Busia and Kimwanyi Zones. The enrolment of participants into the study was voluntary and only after written informed consent is sought from the participant. The participants had the right to withdraw from the study at any time. Identification numbers instead of names of the respondents were used during the research and the data collected were treated with the utmost confidentiality

## RESULTS

359 people participated in the study giving a response rate of 92.05%. The majority of the participants 89.14% were between 18 to 35 years. 243 (67.69%) of the participants were male while 116 (32.31%) were female. 112 (31.2%) of the respondents were Katanga residents while 247 (68.8%) were Makerere University students (Table 1).

**Table 1.**
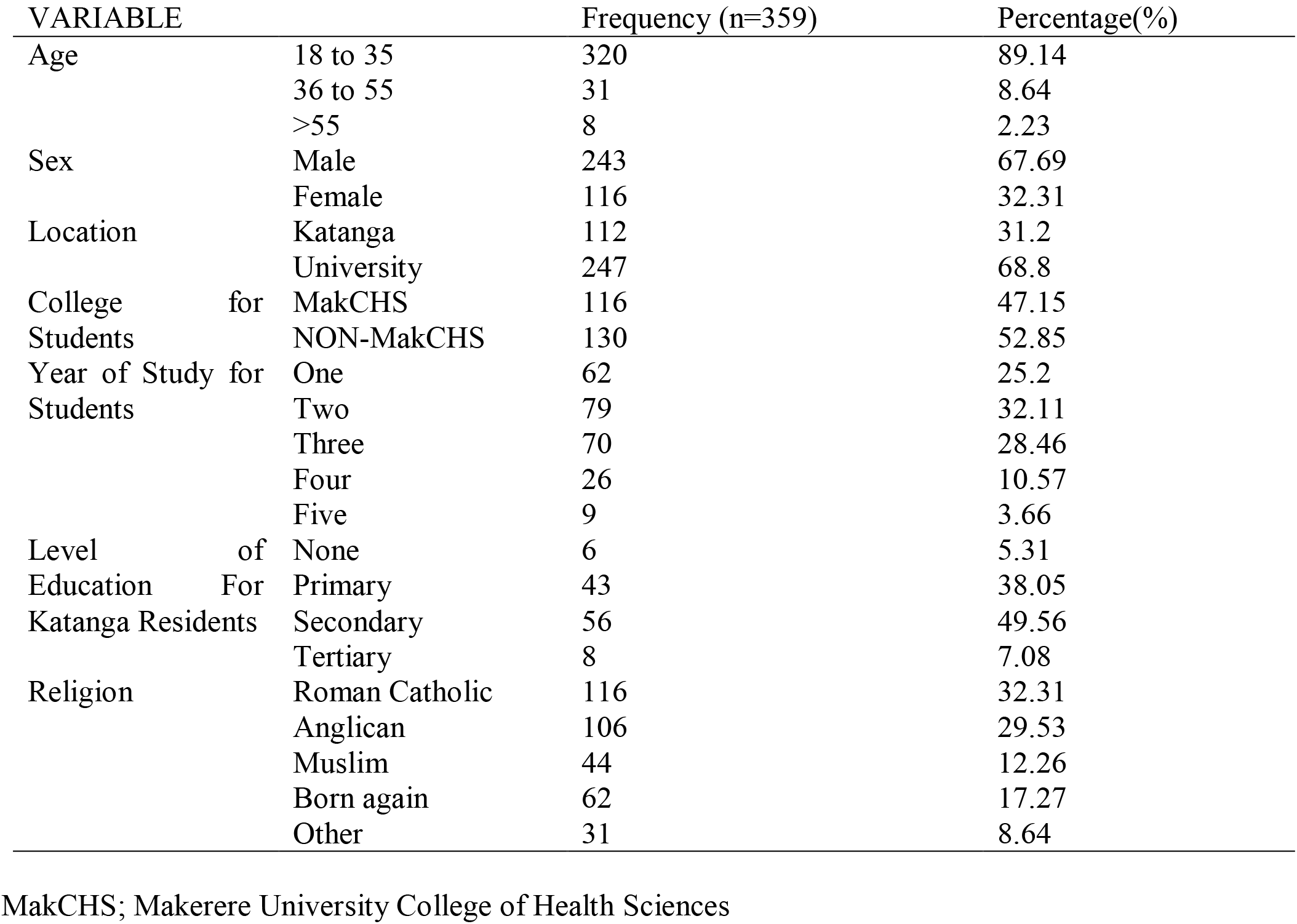
Showing the social demographic Characteristics

## KNOWLEDGE

Only 8.4% of the participants had good knowledge of hand hygiene. All these were young adults (18 to 35 years). All participants above 35 years of age had poor Knowledge of hand hygiene. 11.7% of the university students had good knowledge compared to 0.9% of the Katanga residents. 22.2% of year five students had good knowledge compared to 3.2% of the year students (Table 2). On multivariate analysis, University students were 6.3 times (OR: 6.3, 95%C1: (2.1 – 18.5), P=0.001) more likely to have good hand hygiene knowledge than Katanga residents. There was no significant difference in knowledge on hand hygiene between medical and no medical students. The Spearman’s correlation coefficient between the year of study for university students and the knowledge on hand hygiene was 0.1720, showing a poor positive relationship. The religion, age, and sex of participants did not affect the level of knowledge on hand hygiene.

**Table 2.**
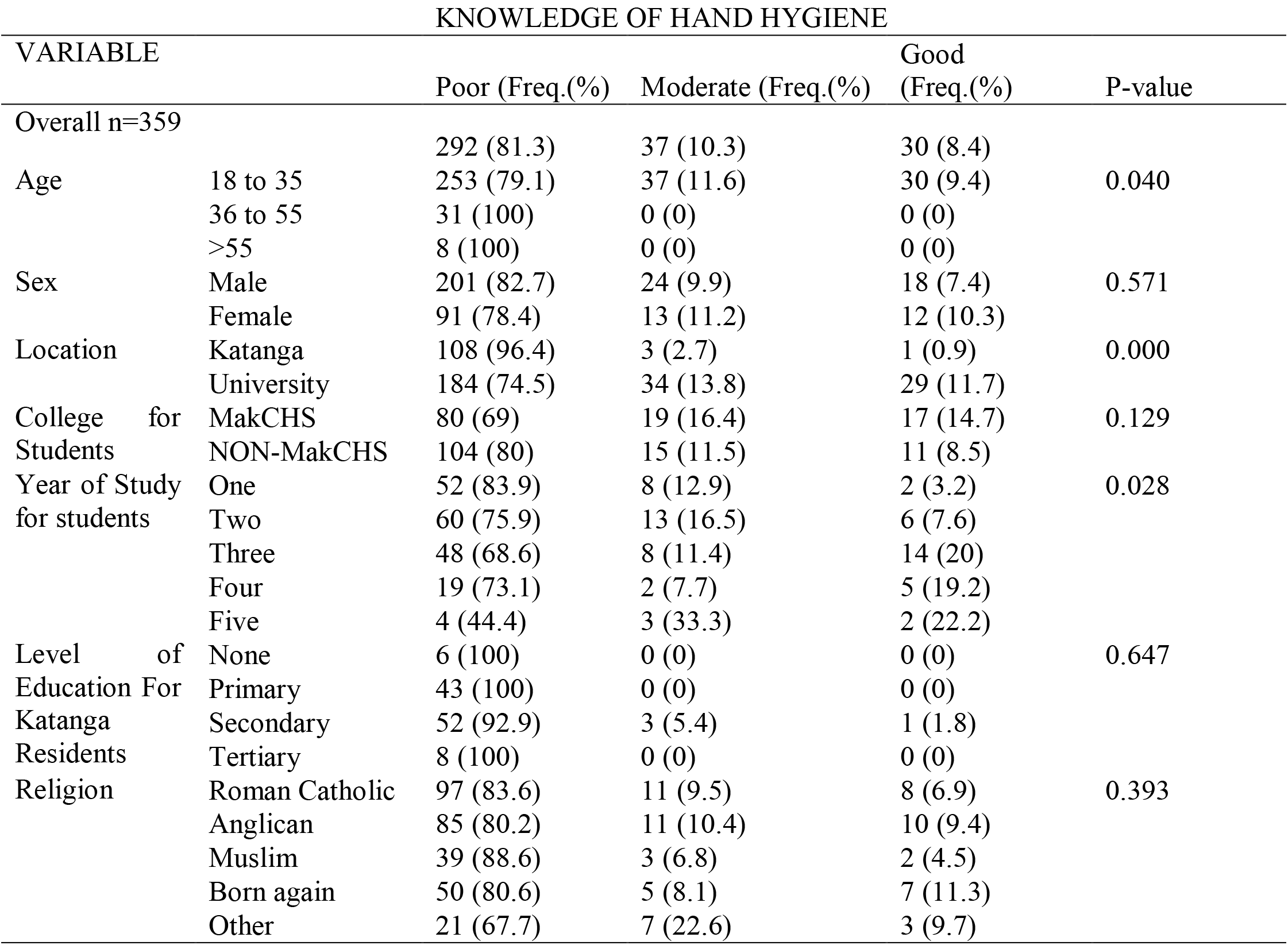
Showing the Knowledge of participants on Hand hygiene

227(63%) of the participants had received prior teaching on hand hygiene and 49% of them had been taught by a health worker. The highest percentage of those trained, 48% had received this training more than 3 months ago. 23.8% of those with teaching on hand hygiene had moderate to good knowledge compared to 9.8 of those that have no training on hand hygiene

Receiving teaching on hand hygiene increased knowledge by 2.9 times (OR: 2.9, 95%CI: (1.5 – 5.5), P = 0.002). How long ago the teaching was delivered did not affect the level of knowledge and neither did the person who delivered the teaching. University students were 3.1 times (OR: 3.1, 95%C1: (2.0 – 5.0), P = 0.000) more likely to have had a teaching on hand hygiene than Katanga residents.

Social media was the most common source of information about hand hygiene as a prevention measure for COVID 19 followed by television, 38.7%, and 28.1% respectively. The commonest source of information among Katanga residents was television (51.8%) followed by radio (25.9%) while among university students, the commonest source of information was social media (52.6%). However, the source of information did not affect the level of knowledge of hand hygiene.

## ATTITUDE

Overall 29.0% of the participants had a good attitude to hand hygiene. The biggest percentage, 50.1% had a moderate attitude to hand hygiene (Table 3). Multivariate analysis showed that Katanga residents were 3.6 times (OR: 3.6, 95%CI: (1.5 – 8.4), P = 0.003) more likely to have a good attitude to hand hygiene than Makerere University students. The college of students and the study year did not affect their attitude to hand hygiene. Although bivariate analysis showed that participants that had been taught hand hygiene prior were 1.8 times (OR: 1.8, 95%C1: (1.1 – 3.0), P = 0.024) more likely to have good attitude to hand hygiene, there was a poor positive relationship between knowledge of the participants and their attitude to hand hygiene due to a spearman’s correlation coefficient of 0.0734.

**Table 3.**
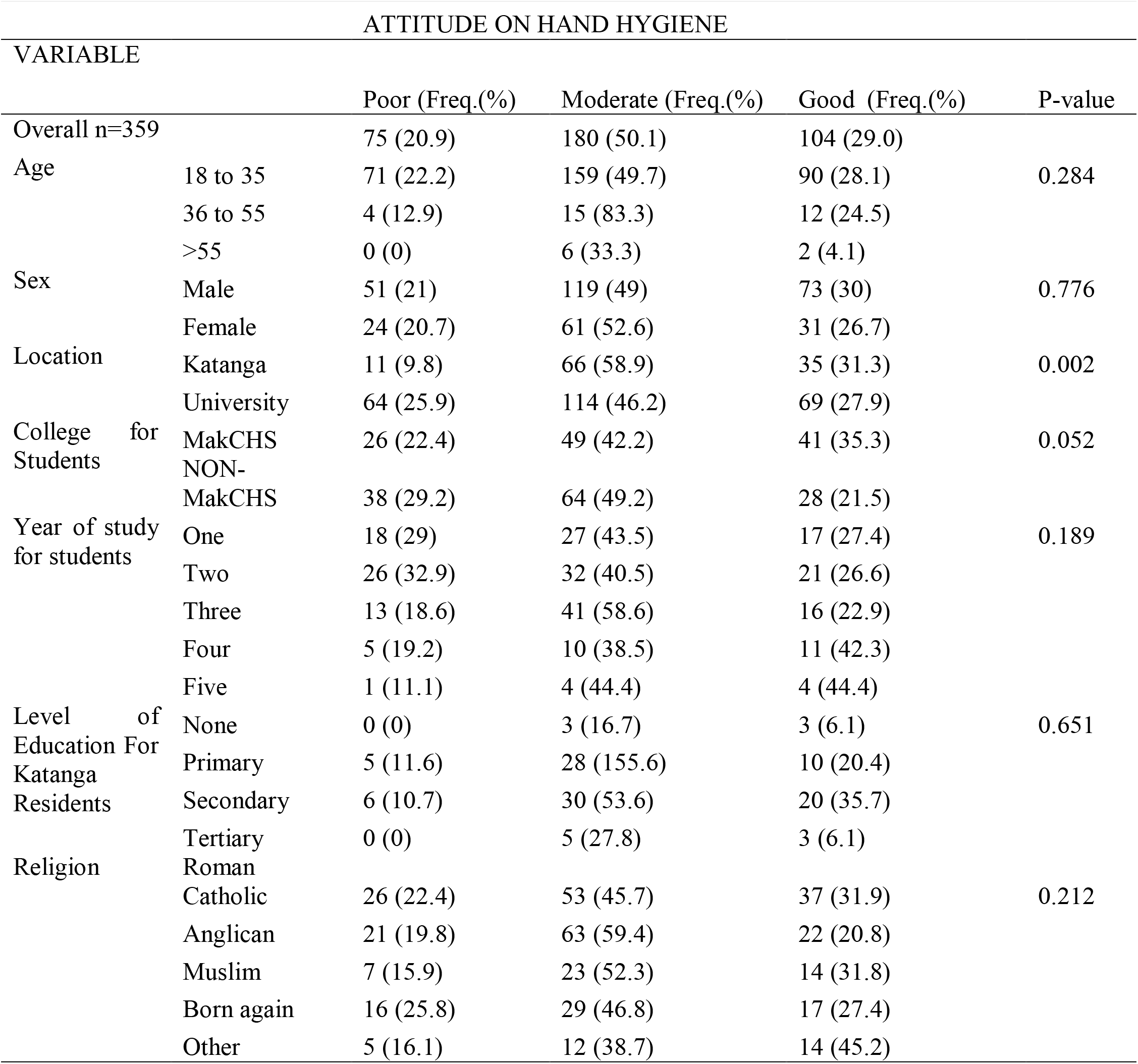
Showing the attitude of participants to hand hygiene

## PRACTICE

138 (38.4%) of the participants were still greeting by handshaking while 149 (41.5%) of the participants greeted with only facial expressions. Only 70 (19.6%) were able to demonstrate all the seven steps of handwashing while 144 (40.2%) demonstrated less than three handwashing steps. Knowledge and attitude of participants on hand hygiene did not affect the way they were greeting in the era of COVID 19 at the time of the study. Participants who demonstrated all the seven steps of handwashing were 9.2 times (OR: 9.2, 95%C1: (4.3 – 20.0), P = 0.000) likely to have been taught handwashing than those who demonstrated less than three steps or demonstrated handwashing without soap use of soap. This had a Spearman’s correlation was 0.3630 showing a fair positive relationship.

## BARRIERS

The biggest barrier to hand hygiene was lack of soap, detergents, alcohol-based hand rub, or antiseptic, reported by 211 (60.1%) of the participants. The most common barrier among university students was lack of soap and or antiseptics, 66.4% while negligence was the most common barrier in Katanga residents, 59.6%. The largest percentage of the participants 62.9% reported having more than three barriers to hand hygiene, (Table 4). University students were 5.3 times (OR: 5.3, 95%C1: (1.6 – 17.6), P = 0.006) more likely to have many barriers to hand hygiene than Katanga residents

**Table 4.**
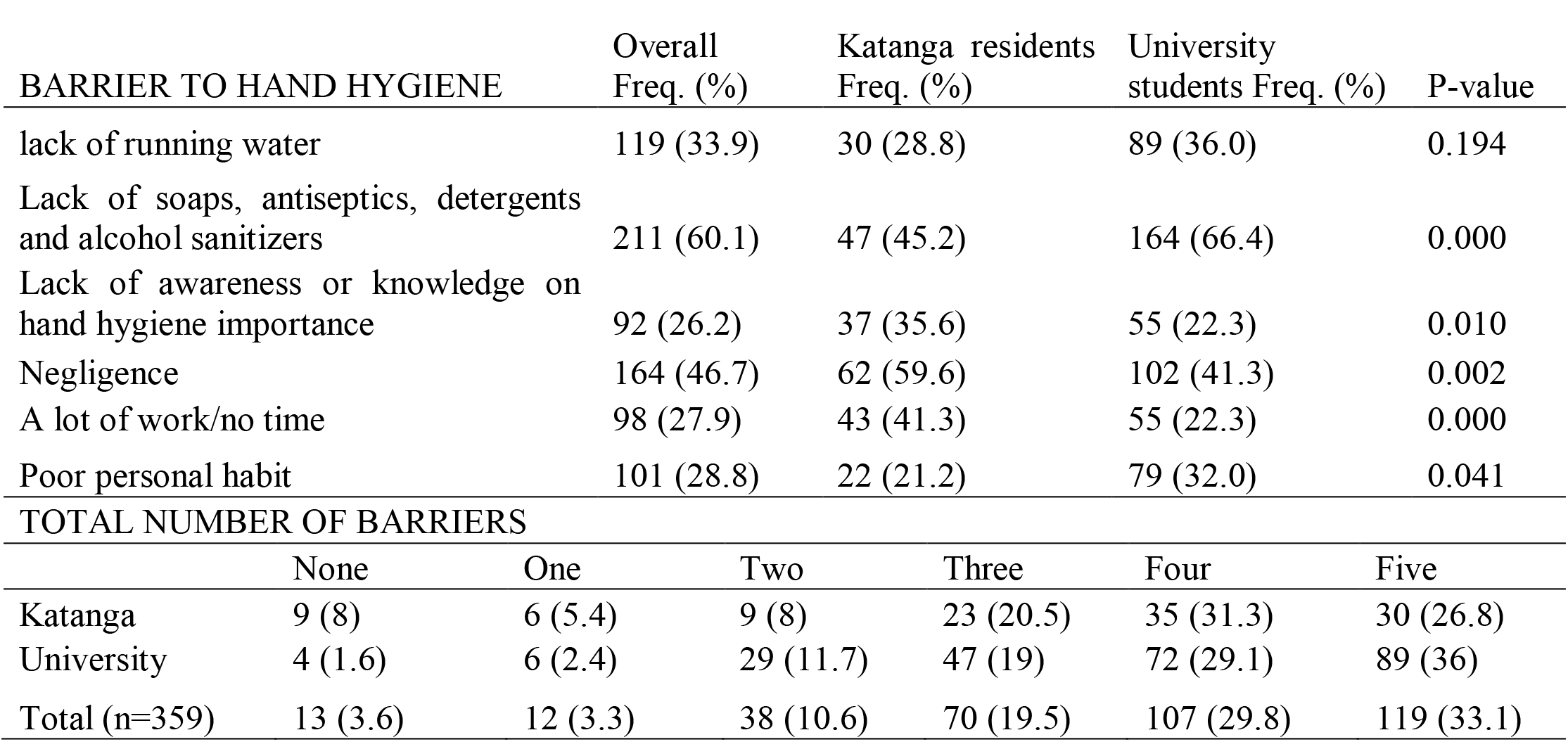
Showing the different barriers to proper hand hygiene

| BARRIER TO HAND HYGIENE | Overall<br>Freq. (%) | Katanga residents<br>Freq. (%) | University<br>students Freq. (%) | P-value |  |  |
| --- | --- | --- | --- | --- | --- | --- |
| lack of running water | 119 (33.9) | 30 (28.8) | 89 (36.0) | 0.194 |  |  |
| Lack of soaps, antiseptics, detergents<br>and alcohol sanitizers | 211 (60.1) | 47 (45.2) | 164 (66.4) | 0.000 |  |  |
| Lack of awareness or knowledge on<br>hand hygiene importance | 92 (26.2) | 37 (35.6) | 55 (22.3) | 0.010 |  |  |
| Negligence | 164 (46.7) | 62 (59.6) | 102 (41.3) | 0.002 |  |  |
| A lot of work/no time | 98 (27.9) | 43 (41.3) | 55 (22.3) | 0.000 |  |  |
| Poor personal habit | 101 (28.8) | 22 (21.2) | 79 (32.0) | 0.041 |  |  |
| TOTAL NUMBER OF BARRIERS |  |  |  |  |  |  |
|  | None | One | Two | Three | Four | Five |
| Katanga | 9 (8) | 6 (5.4) | 9 (8) | 23 (20.5) | 35 (31.3) | 30 (26.8) |
| University | 4 (1.6) | 6 (2.4) | 29 (11.7) | 47 (19) | 72 (29.1) | 89 (36) |
| Total (n=359) | 13 (3.6) | 12 (3.3) | 38 (10.6) | 70 (19.5) | 107 (29.8) | 119 (33.1) |

## DISCUSSION

COVID 19, is a global pandemic with a high transmission rate (27). One of the best measures to address the spread of COVID 19 is adherence to proper hand hygiene practice (14). It is therefore important to understand the knowledge, attitude, and practice of high-risk areas to hand hygiene. Together with establishing the existing barrier to hand hygiene, solutions can be formulated on proper infection prevention to limit the spread of COVID 19 in the communities.

Overall, only 8.4% of the participants had good knowledge of hand hygiene, a finding that is similar to a study in India and among medical students at Kampala International University (28,29). 11.7% of the university students had good knowledge compared to 0.9% of the Katanga residents. 63% of the participants had ever received teaching on hand hygiene, this left 37% of the participants without prior teaching on hand hygiene. This demonstrated the need for health education on hand hygiene to address this gap. 23.8% of those with teaching on hand hygiene had moderate to good knowledge compared to 9.8% of those that had no training on hand hygiene This knowledge was not affected by who delivered the teaching and or when it was delivered. This concluded that any person with hand hygiene knowledge, is positioned to deliver information to another individual and this should be promoted. The commonest sources of information on hand hygiene as a control measure for the spread of COVID 19 were Social media (38.7%), Television (28.1%), radio (11.4%), etc., in descending order. This shows the importance of these forms of communication in connecting with communities (30,31) and this should be used to promote hand hygiene in this pandemic.

The study found that 29.0% of the participants had a good attitude and 50.1% had moderate attitudes to hand hygiene which is similar to a study among Kampala International University medical students who were found to have a positive attitude to hand hygiene (29). A high number of Makerere University students 25.9% were found to have a poor attitude to hand hygiene compared to Katanga slum residents 9.8%. This can be explained by the poor positive relationship between attitude and hand hygiene knowledge.

All the social demographic characteristics did not affect the knowledge and attitude of participants to hand hygiene except their location, a finding that is similar to a study among Chinese adults (32). Amidst the growing worldwide incidence and the fact that Uganda had already registered the index case of COVID 19 (33). Our study established that 38.4 % of the participants were still greeting by handshaking and/or hugging. This showed a poor hand hygiene practice among these participants and subsequent high risk of community transmission of COVID 19. Only 19.6% of the participants were able to demonstrate the 7 steps of handwashing. Participants who had been taught had hygiene were 9.2 times more likely to demonstrate all the seven steps than their counterparts, showing the importance of hand hygiene education and promotion as a means of improving its proper practice (21).

The commonest barriers to hand hygiene were lack of soap, lack of soaps, antiseptics, detergents and alcohol sanitizers (, lack of running water and negligence findings that are no different from a study by Muiru (29) and a study by Al-Naggar which showed that laziness was the main barrier (34). The majority of the patients had more than three barriers to hand hygiene. This is a very big public health problem that limits proper hand hygiene and which is the single most effective protective measure against COVID 19 (11–13), and it needs to be addressed.

## CONCLUSION AND RECOMMENDATIONS

The majority of the participants had poor knowledge of hand hygiene. However, many of the participants had a moderate attitude to hand hygiene. The practice of hand hygiene was found to be low and the majority of the participants faced more than three barriers to hand hygiene with the most common barrier being lack of soap, detergent, or antiseptic. The knowledge and attitude to handwashing were found to be influenced by the location of the participants. The study also established that 37% of the participants had never received teaching on hand hygiene. These findings show that a lot has to be done if had hygiene is to be effective as a measure to prevent the community spread of COVID 19.

Based on this study, more public health involvement is needed to promote hand hygiene. This can involve health education campaigns on social media, television, and radio stations since they are the safe ways of communication in times of such a pandemic like COVID 19. The hand hygiene health education can further be scaled to community campaigns when the pandemic is over. We recommend that equipped hand washing facilities are set up in communities and institutions, this should be done to address the several physical barriers that affect proper hand hygiene. Studies should do be carried out to find solutions to the solutions.

## Data Availability

The dataset presented in this study can be found on FigShare

https://figshare.com/articles/MAY_8TH_data_sheet_KAP_COVID_19_-_for_analysis_xlsx/12435341

## ACKNOWLEDGMENTS

We are grateful to residents of Katanga Slum and students of Makerere University for participating in the study. We acknowledge the following people; Keneth Kato Agaba, Julius Musisi, Kevin Adongo, Laurita Nakyagaba, Patricia Mwachan, Abraham Byomugabe, Mavol Tukwataniise, Ivan Ongebo, and John Muhenda, for participating in data collection. We thank Prof. Sarah Kiguli and Dr. Margrete Lubwama for supervising us in the study, Ronald Olum for his guidance in analysis and manuscript writing and Prof. Charles Ibingira through the office of the principal Makerere University college of health sciences for authorizing funding the research process.

## DATA AVAILABILITY STATEMENT

The dataset presented in this study can be found on FigShare at, https://figshare.com/articles/MAY_8TH_data_sheet_KAP_COVID_19_-_for_analysis_xlsx/12435341

